# Inflammatory Bowel Disease in patients with Primary Sclerosing Cholangitis: a distinct form of colitis

**DOI:** 10.1101/2024.07.23.24310848

**Authors:** Amber Bangma, Paola Pibiri, Shiqiang Sun, Sofie De Jong, Emilia Bigaeva, Johannes R. Björk, Naomi Karmi, Gwenny M.P.J. Verstappen, Frans G.M. Kroese, Gursah Kats-Ugurlu, Marcela A. Hermoso, Arnau Vich Vila, Monique G.P. van der Wijst, Klaas N. Faber, Rinse K. Weersma, Werna T.C. Uniken Venema, Eleonora A.M. Festen

**Affiliations:** Department of Gastroenterology and Hepatology, University of Groningen, University Medical Center Groningen, Groningen, The Netherlands; Department of Genetics, University of Groningen, University Medical Center Groningen, Groningen, The Netherlands; Department of Pathology, University of Groningen, University Medical Center Groningen, Groningen, The Netherlands; Department of Rheumatology and Clinical Immunology, University of Groningen, University Medical Center Groningen, Groningen, The Netherlands; Universidad de Chile, Biomedical Sciences Institute, Faculty of Medicine, Immunology Program, Innate Immunity Laboratory, Santiago, Chile

**Author notes:** **CORRESPONDING AUTHOR**: Eleonora A.M. Festen, Department of Gastroenterology and Hepatology, University Medical Center Groningen, PO Box 30.001, Hanzeplein 1, 9700 RB Groningen, the Netherlands. Authors share first authorship (Amber Bangma, Paola Pibiri, Shiqiang Sun). Authors share last authorship (Werna T.C. Uniken Venema, Eleonora A.M. Festen).

**Keywords:** PSC-IBD, UC, DUOX2, auto-immune diseases, inflammation

## Abstract

**Background and aims:** Primary sclerosing cholangitis (PSC) is an inflammatory disorder of the bile ducts, often accompanied by inflammatory bowel disease (PSC-IBD). Substantial differences in clinical presentation are observed between PSC-IBD and ulcerative colitis (UC). In this study we aim to find distinct pathomechanisms for PSC-IBD using single-cell mRNA sequencing.

**Methods:** Forty-seven colonic mucosal biopsies of PSC-IBD (n=24), UC (n=18) (where possible matched inflamed (I) non-inflamed (NI)), and non-IBD subjects (n=5) were collected and dissociated. Library preparation and processing for sequencing was followed by differential abundance, differential expression, and cell-cell-interaction analyses. Stainings for DUOX2 and HLA-DR were applied on tissue sections.

**Results:** In total, 71,798 cells comprised 54 distinct cell types, including a new cell type: the DUOX2+ enterocyte, mainly present in inflamed colon and acting as antigen presenting cells (HLA-DR+). Stem cells exhibited increased abundances in PSC-NI but not in UC-NI. Additionally, we found distinct gene expression profiles in PSC and UC which were related to inflammation: while mainly inflammatory monocytes were activated in PSC inflammation, and inflammatory fibroblasts were activated in UC inflammation.

**Conclusion:** Our study found a new cell type, the DUOX2+ enterocyte that is primarily present in inflamed conditions, and based on their expression profile, potentially perform antigen presentation. Moreover, we highlight that PSC-IBD, but not UC, is characterized by the activation of inflammatory HLA-DRB1+ monocytes, which are likely involved in the activation of CD4+ T cells. Notably, we observed an increased abundance of stem cells in non-inflamed PSC-IBD, possibly linked to the elevated risk of colorectal cancer in PSC-IBD.

**What is known on the subject?:** Inflammatory Bowel Disease (IBD) presents in 60–80% of primary sclerosis cholangitis (PSC) patients, termed PSC-IBD. However, the clinical presentation is distinct: patients have 10 times increased risk of colorectal carcinoma as compared to ulcerative colitis (UC) patients, and the conventional IBD treatment regimen is not always effective. The underlying genetic predisposition is also different from general IBD, where PSC-IBD shows a very strong risk association in the HLA-DRB1 region, generally a hallmark of auto-immune disease.

**What this study adds:** This study uncovers that PSC-IBD has more autoimmune and UC more inflammatory characteristics. This is supported by the interaction between inflammatory monocytes and CD4+/CD8+ T cells through HLA-DR in PSC-IBD. PSC-IBD patients have pronounced increased activity of inflammatory monocytes, not seen in UC. We find increased stem cell abundance in PSC-IBD. In addition, we show cellular-molecular mechanisms potentially driving the increased risk of carcinoma in PSC-IBD patients offering valuable foundations for targeted therapeutic strategies. In addition, we determined the new DUOX2+ enterocyte.

**How this research might affect research, practice, or policy:** This study uncovers that PSC-IBD has more autoimmune characteristics whereas UC is an immune mediated inflammatory disease. This is supported by the specific interaction between inflammatory monocytes and CD4+/CD8+ T cells through HLA-DR in PSC-IBD, while in UC DUOX2+ enterocytes and inflammatory monocytes communicate with innate immune cells, Tregs and CD4+ T cells. Our findings may form new treatment targets for PSC-IBD and IBD in general.

## Introduction

Primary Sclerosing Cholangitis (PSC) is an inflammatory disorder impacting the bile ducts, occurring in approximately 4-20 per 100,000 individuals and showing a predilection for men over women[1–3]. The majority of PSC patients need liver transplantation within 10-20 years from diagnosis, with disease recurrence in 30-50% of patients within 10 years post-transplantation. As of now, there is no cure for PSC[4–6].

In 60-80% of cases, PSC patients concurrently experience inflammatory bowel disease (IBD). The clinical presentation of PSC-IBD shares similarities with ulcerative colitis (UC), but distinctive features include a preferential involvement of the right side of the colon, in contrast to the typical left-sided involvement in UC. Notably, individuals with PSC-IBD face an elevated risk, up to five times higher, of developing colorectal carcinoma (CRC) compared to UC patients (2% at 10 years after diagnosis[7]), necessitating annual surveillance endoscopies[4,8,9]. While current treatment protocols for PSC-IBD mirror those for regular IBD, studies indicate a diminished response to anti-tumour necrosis factor alpha (anti-TNFa) therapy in PSC-IBD compared to conventional IBD[10,11]. These differences between PSC-IBD and UC are hypothesized to originate from variations in the underlying mechanisms of the two forms of IBD.

Differences in the genetic risk background between PSC-IBD and IBD support this hypothesis: where UC and Crohn’s disease (CD) have a genetic correlation of r = 0.56, the genetic correlations between UC and PSC-IBD (r = 0.29) and between CD and PSC-IBD (r = 0.04) are considerably weaker. In fact, the PSC-IBD genetic risk locus at *HLA-DRB1* resembles patterns of auto-immune diseases such as coeliac disease and type 1 diabetes, suggesting that PSC-IBD pathophysiology might rely on presentation of an unknown antigen, with lymphocyte activation as a result[12]. Although the exact mechanism remains unclear, colonic inflammation is suggested to be relevant for progression of bile duct disease in PSC, since colectomy improves outcomes after liver transplantation[13]. An explanation for the increased CRC risk in PSC patients is currently lacking.

We hypothesize that a distinct disease process underlies PSC-IBD in comparison to UC, contributing to divergent phenotypes and varied disease outcomes. Here, we investigate 47 inflamed and non-inflamed colonic biopsies of PSC-IBD, UC and non-IBD patients using single-cell mRNA-sequencing (scRNA-seq) and describe commonalities and specificities in cell composition together with signalling pathways between PSC-IBD and UC. Obtaining a basic understanding of the pathomechanisms underlying PSC-IBD is a crucial first step towards specific treatment regimens for PSC-IBD.

## Materials and methods

### Sample collection and selection

A total of 47 samples were collected from the colon of subjects with either PSC-IBD, UC or without IBD during a routine colonoscopy at the University Medical Center Groningen (Groningen, the Netherlands). The diagnosis IBD was based on typical endoscopic findings confirmed by histopathological evidence. The diagnosis PSC was based on cholestatic blood levels in combination with typical radiological findings, after ruling out other possible causes for bile duct sclerosis. “Healthy control” (HC) samples were collected from patients without IBD, who underwent endoscopy for various reasons, such as rectal bleeding, check-up after polyp removal or abdominal discomfort. The absence of IBD was histologically confirmed by a trained pathologist. One “HC” sample was excluded since the patient’s diagnosis was revised to IBD at clinical re-evaluation.

Additional inclusion criteria for PSC-IBD and UC participants were no history of liver transplantation, and no isolated sigmoid involvement of the disease. Consent of all participants was obtained through Gastrointestinal Expression profiles in Intestinal Disease (GE-ID) (NL58808.042.16) and/or Parelsnoer IRB (NL24572.018.08). At the time of endoscopy, three biopsies per participant were collected in RPMI with 10% FCS at 4°C, and cryopreserved into ice cold freezing medium (FCS with 10% DMSO) within 30 minutes after harvesting. At least one biopsy from the same location as the cryopreserved biopsies was evaluated by a pathologist to examine inflammatory activity. Based on the histological evaluation, biopsies were classified as actively inflamed or non-inflamed.

### Sample dissociation

Three gut mucosal biopsies per sample were thawed as described before[14,15], and subsequently dissociated using the ‘one-step collagenase’ protocol[15]. In short, biopsies were washed with ice cold PBS with 10% FCS for 15 minutes. Then they were digested with collagenase IV and DNAse II for 30 minutes in RMPI with 10% FCS at 37°C, while being pipette-strained at t=10 minutes and t=30 minutes. Finally, the biopsies were shortly incubated in TrypLE, filtered over a 70 μm filter and washed using cold PBS with 1% BSA.

### Cell hashing

Cells were hash tagged using Totalseq A antibody hashtag 1 to 4 (Biolegend), according to the manufacturer’s protocol. These hashtags were later used to demultiplex pooled samples.

### Viability assessment and cell counting

Viability and counting before loading were assessed using trypan blue staining and counting on a hemocytometer. At viability <60%, cells were washed once more, and when visible clumps were present, cells were filtered once more over a 70 μm filter.

### Library preparation

The 10X V3 chip (B) with V3 reagents were used for library preparation. Four samples were pooled per 10X lane, aiming for a total recovery of 12,000 cells per lane. Per chip, 2 channels were loaded. cDNA library prep was done according to the 10X manual CG000185 Rev D, and hashtag libraries were prepared using the manufacturer’s protocol [16]. Libraries were indexed using 10X PN-120262 Chromium i7 Multiplex Kit.

### Library quality control (QC)

Libraries were checked for concentration and peak-size distribution, using an Agilent 4200 TapeStation. All cDNA libraries featured concentration > 20 ng/ul.

### Sequencing and data preparation

Sequencing was performed by BGI (Hong Kong) on an MGI2000 sequencer, featuring 100bp paired-end reads (90k reads per cell). Demultiplexing was performed at BGI, and subsequently, Cellranger v3.1.0[17] was used to perform filtering, barcode counting, and alignment, using the human GRCh38-3.0.0 genome as a reference. Detection of possible doublets was primarily performed using hashtag assignment, with additional assignment using Souporcell (v2.0)[18].

Seurat[19] v2.3.0 was used for data QC and analysis. Cells were filtered for >200 genes and <60% mitochondrial reads per cell. After integration, RNA counts were normalized with log-normalization through Seurat. Next, the first 30 principal components (based on the elbow plot) were used in the RunPCA function in Seurat for Principal Component Analysis (PCA). Finally, the FindNeighbors and FindClusters functions (with default settings) were utilized to detect cell clusters.

### Cell type annotation

As a first step, all high-quality cells were divided into three compartments: epithelial, stromal, and immune. Specifically, the Seurat object was used to calculate the average expression of compartment markers for each cluster, using the function AverageExpression to identify epithelial cells (EPCAM >=4), stromal cells (THY1 >= 0.3, SOX10 >=2, MADCAM1 >=1), and immune cells (CD27 >=0.3, PTPRC >=3). The immune compartment was further filtered for <20% mitochondrial reads per cell. Next, each compartment was re-clustered and annotated separately using an automated reference-mapping algorithm (azimuth.hubmapconsortium.org) using a UC full biopsy scRNAseq dataset[20] as a reference set on the integrated (SCT) assay. This method relies on a reference single-cell dataset, where cells in the query dataset are labelled based on their closest projected reference cell (cell-to-cell approach). Lastly, biologically relevant cell types were manually annotated through marker gene clustering. Specifically, dual oxidase 2-expressing (DUOX2+) enterocytes (*DUOX2*>1) were annotated within the epithelial compartment; and IgA (*IGHA1*>115), IgG (*IGHG1*>90), IgM (*IGHM*>800), and Ig-negative plasma cells were annotated within the immune cell compartment.

### Differential abundance analysis

To perform differential cell abundance analysis, the Bayesian Multinomial Logistic-Normal Linear Regression model Pibble for compositional data (R package Fido v1.0.4[21]) was used. Because the Pibble model uses Bayesian inference for parameter estimation, statistical significance is expressed as the credible interval; for example, a cell type can be deemed significantly differentially abundant if 95% of its credible interval does not contain zero. Furthermore, as the Pibble model estimates the correlation structure between all included cell types (after adjusting for covariates if any), model inference is only done once.

### Differential gene expression and pathways

Differentially expressed genes were defined as genes expressed in at least 10% of either of the two comparison groups and with a minimum of 0.25-fold change in gene expression. For differential gene expression Seurat (v4.3.0.1) with the built-in ‘MAST’ method (v1.26.0) was used. All p-values were Bonferroni corrected for the number of tested genes. Pathways were then generated using the R package enrichR (v3.2) with the GO biological process 2018 database using DE genes as input[22].

A list of significantly upregulated genes (marker genes) for each cell type was generated using the above-described method, comparing the respective cell to all other cells within the cell type’s compartment (e.g. DUOX2+ enterocytes were compared to all other epithelial cells). With the list of marker genes as input, a list of marker pathways was then generated for “DUOX2+ enterocytes” and “enterocytes” accordingly.

Differentially expression of oncogenes was assessed using the above-described method, using pancancer and colorectal cancer onco(suppression)genes as input[23].

### Cell-cell interaction analysis

Cell-cell interaction analysis was performed for Inflammatory Monocytes and DUOX2 Enterocytes in relation to other cell types with the R package CellChat (v.1.4.0), using a public build-in database on ligand-receptor interactions[24].

### Heritability analysis

Heritability enrichment analysis was performed using the CELL-type Expression-specific integration for Complex Traits (CELLECT) v1.3.0 toolkit[25], which is designed to identify likely etiologic cell types underlying complex traits. This toolkit applies stratified LD score regression (S-LDSC)[22] to determine cell types with genetic associations significantly enriched for the disease or trait of interest. Standard settings were applied in the implementation of CELLECT, including the exclusion of genetically complex regions like the HLA locus prior to analysis.

For heritability enrichment analysis for PSC specifically expressed genes, genome-wide association study (GWAS) summary statistics of PSC from a comprehensive PSC GWAS[26]. was used as the genetic summary statistics of interest, and CELL-type EXpression-specificity (CELLEX) analysis[25] was performed to generate cell type specific gene scores.

For heritability enrichment analysis for differentially expressed genes between PSC-I and PSC-NI, we performed MAST v1.14.0, a two-part, generalized linear model with a logistic regression component for the discrete process (i.e., a gene is expressed or not) and linear regression component for the continuous process (i.e., the expression level)[27] to generate differential gene scores. Inflammation within PSC, sex, age, mitochondrial percentage, and the number of genes detected per cell[20,27] were included as fixed effect variables and individual as a random effect to control for pseudoreplication bias[28]. The Benjamini-Hochberg correction was used to control for multiple testing across all cell types. GWAS summary statistics for PSC from the previously mentioned publication[26] was used as the genetic summary statistics of interest.

For both types of heritability enrichment analysis (PSC specifically expressed genes and differentially expressed genes between PSC-I and PSC-NI), Benjamini-Hochberg correction was applied to control for the total number of tests across all cell types.

### Immunohistochemistry

To confirm the existence of DUOX2+ enterocytes and the expression of HLA-DR at the protein level, we performed DUOX2 and HLA-DR immunohistochemical staining of inflamed and non-inflamed colonic mucosa. Sequential 4µm slides from resected colonic paraffin-embedded tissue were deparaffinized, followed by antigen retrieval with a Citrate buffer pH6 and blocking of endogenous peroxidase with 0.3% hydrogen peroxidase (Sigma-Aldrich). Next, the slides were incubated with PBS/ 1% BSA (Sigma-Aldrich) for 30 min, followed by incubation with a primary antibody – DUOX2 (1:100 dilution, cat. PA5-97677, Thermo Fisher) or HLA-DR (1:1000 dilution, mouse monoclonal DA6.147, Abcam) for 1 hour in the dark. Appropriate secondary and tertiary horseradish-peroxidase labelled antibodies were used for both stainings and diluted with a 1:50 ratio in staining solution (PBS with 1% BSA and 1% human serum). To complete the staining, a solution of DAB (Sigma-Aldrich) and 0.03% peroxidase was used, followed by a counterstaining of haematoxylin and mounting with Eukitt (Sigma-Aldrich). Slides were digitally scanned using a Nanozoomer (Hamamatsu). Images were taken at 5x and 40x magnification using NDP view 2 software. A pathologist specialized in gastrointestinal disease was consulted to assess the staining.

## Results

### Primary results after QC

In this study we applied scRNA-seq to 47 gut biopsy samples from 24 PSC-IBD (8 inflamed (PSC-IBD-I), 16 non-inflamed samples (PSC-IBD-NI)), 18 UC (8 inflamed (UC-I), and 10 non-inflamed samples (UC-NI)), and 5 healthy control (HC) samples. The donor and treatment characteristics of these groups were comparable, except for a higher percentage of biopsies from male patients (p=0.04) and patients using of ursodeoxycholic acid in the PSC-IBD group (p=0.001) (in line with epidemiology / standard care of disease). Furthermore, UC biopsies were significantly more often taken from patients exposed to biologicals (0.0074).

After quality control and filtering, we identified 71,798 cells from 47 samples. These cells represented 54 cell types: 25 immune, 16 epithelial, 12 stromal and 1 glia cell type. Marker genes for each cell type within its compartment are available in Supplementary results 1. Figure 1A shows the non-Euclidian distribution of the cells per compartment based on their transcriptional profile in a two-dimensional visualization of multiple principal components (Uniform Manifold Approximation and Projection - UMAP).

**Figure 1.**
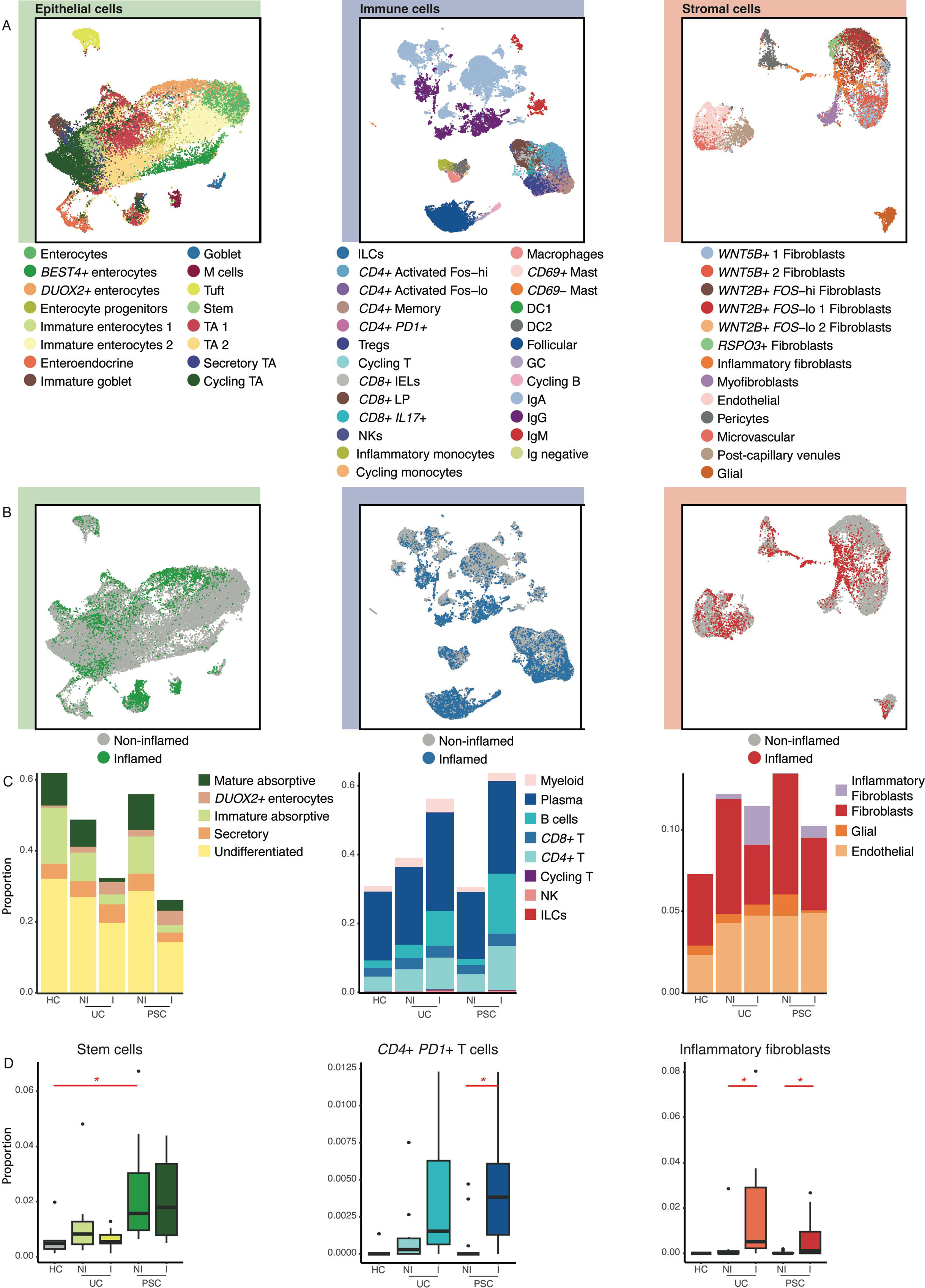
A: UMAP view of 71,798 cells divided into the three compartments (epithelial, immune and stromal) single-cell transcriptomics coloured by annotation cell type. B: UMAP view of the total cells divided into the three compartments (epithelial, immune, and stromal) and coloured by inflammation status. Non-inflamed state is composed of non-inflamed samples of HC, UC and PSC. Inflamed state is composed of inflamed samples of UC and PSC. C: Cell proportion of a combination of cell types as a fraction of the total, here represented within each compartment (epithelial, stromal, and immune). Specification of cell types included in each cell group is available in supplemental results 1. The total of cells is considered within each condition (HC, UC-NI, UCI, PSC-NI, PSC-I). D: Cell proportions of stem cells, CD4+ PD1+ T cells and inflammatory fibroblasts in different states within the entire set (epithelial, immune, and stromal compartment combined).

### DUOX2+ enterocytes, a distinct epithelial cell type involved in antigen presentation in inflammation

In addition to previously reported cell types, we describe dual oxidase 2 (DUOX2)-expressing enterocytes as a new subtype of enterocyte. This enterocyte subtype is marked by the expression of *DUOX2*, its accessory protein dual oxidase maturation factor 2 (*DUOXA2*), and lipocalin-2 (*LCN2*). Staining of paraffin embedded inflamed colonic tissue shows DUOX2^+^ enterocytes present in patches in inflamed tissue, clearly distinguishable from goblet cells that show membrane staining of DUOX2 (Figure 2D), whereas DUOX2^+^ enterocytes show intracellular staining. This cell subtype is distinct from other epithelial cells based on both protein and gene expression. Interestingly, DUOX2^+^ enterocytes are more abundant in inflamed tissue (Figure 2A and Supplementary Results 2).

**Figure 2:**
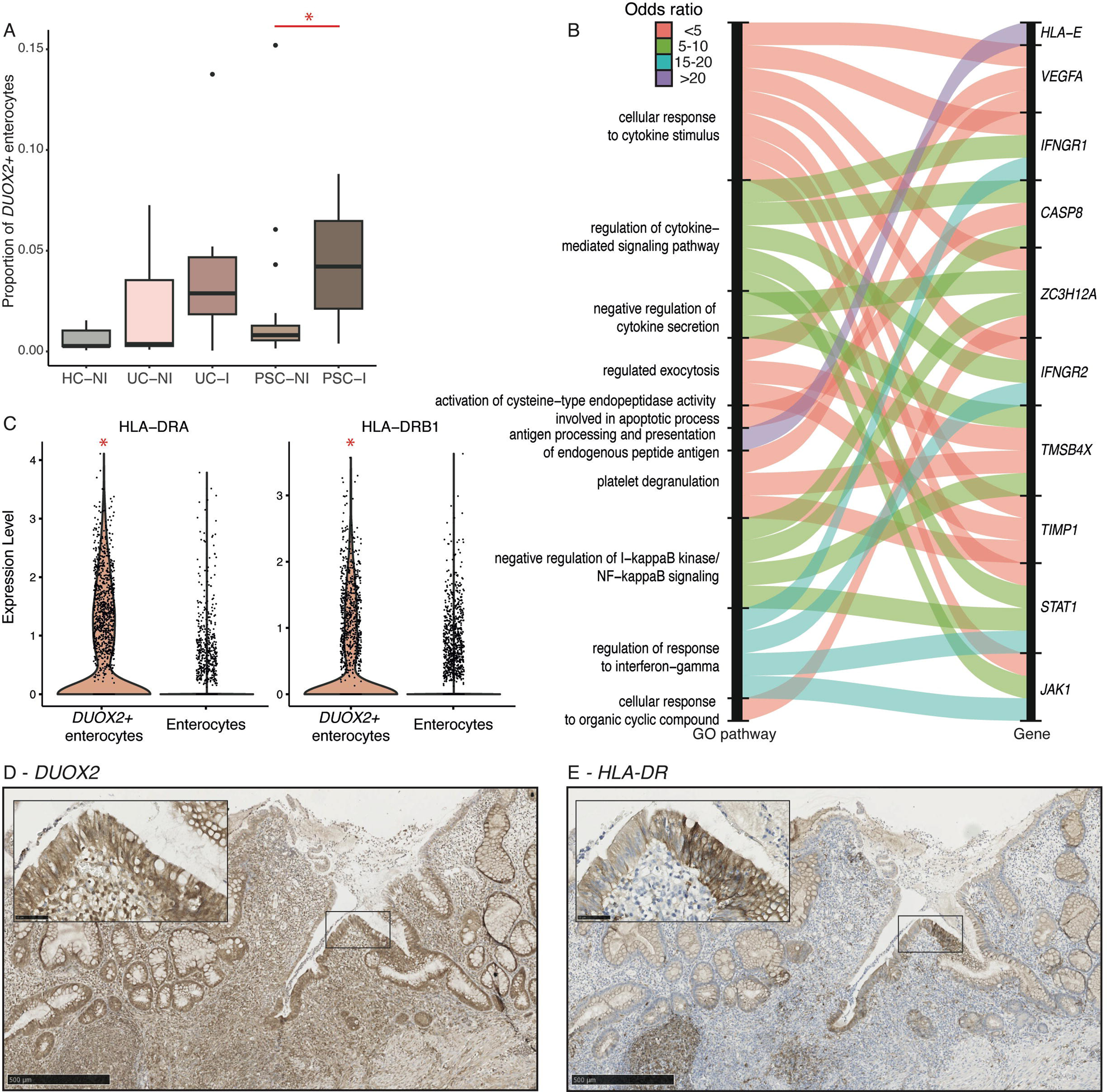
A: Cell proportion of DUOX2+ enterocytes in different states within the entire set (epithelial, immune and stromal compartment combined). Significant differences are denoted with an asterisk. B Alluvial plot representing top 10 most significant marker (GO) pathways of DUOX2+ enterocytes which are not marker pathways for enterocytes and marker genes of these pathways. The colours represent the odds ratio of the marker pathway. C: Expression level of *HLA-DRA* and *HLA-DRB1* in DUOX2+ enterocytes and enterocytes. Marker genes for the given cell types are denoted with an asterisk. D and E: DUOX2 and HLA-DR protein stainings in UC-I colon tissue.

To define how DUOX2^+^ enterocytes differ from other enterocytes, we analyzed pathway enrichment in DUOX2 vs mature (non-DUOX2) enterocytes. Of the 158 pathways enriched in DUOX2+ enterocytes, 99 were not observed in mature (non-DUOX2) enterocytes (Supplemental results 2). The top three pathways specific to DUOX2^+^ enterocytes are those involved in cellular responses to cytokine stimuli: “cellular response to cytokine stimulus” (adjusted p-value=8.81e-06), “regulation of cytokine-mediated signalling pathway” (adjusted p-value=4.47e-04), and “negative regulation of cytokine secretion” (adjusted p-value=1.10e-03), as shown in Figure 2B. Interestingly, among DUOX2+ marker genes we find the MHC-II encoding genes *HLA-DRA* (adjusted p-value=2.67e-143), *HLA-DRB1* (adjusted p-value=1.66e-77), *HLA-DQB1* (adjusted p-value=4.69e-37), and *HLA-DPA1* (adjusted p-value=7.75e-31) (Figure 2C). Moreover, protein staining shows that DUOX2^+^ enterocytes express HLA-DR on a protein level, supporting the hypothesis that these cells can present antigens to CD4^+^ T cells (Figure 2E). Notably, the HLA-DR protein distribution in DUOX2^+^ cells is specific to inflamed colon tissue and staining is restricted to subepithelial macrophage lamina propria cell infiltrate (Supplementary Figure 1F).

DUOX2^+^ enterocytes seem to behave differently upon inflammation in PSC-IBD compared to UC, as shown by differential gene expression (Supplemental results 4-7). The most significantly upregulated gene in PSC-IBD-I vs PSC-IBD-NI (not upregulated in UC-I), is *HLA-DRB1* (adjusted P-value=8.71e-23), which has previously been linked to PSC-IBD pathogenesis[12]. DUOX2^+^ enterocytes in PSC-IBD-I vs PSC-IBD-NI upregulate interferon-stimulated and inducible genes (resulting in “type I interferon signaling pathway” expression) (adjusted P-value=2.67e-03) and also MHC-II encoding genes (“antigen processing and presentation of exogenous peptide antigen via MHC class II”) (adjusted P-value= 2.24e-03); whereas DUOX2^+^ enterocytes in UC-I upregulate mucin-genes involved in “O-glycan processing” (adjusted P-value= 6.47e-04). The three most significantly enriched pathways in both PSC and UC inflammation in DUOX2^+^ enterocytes involve neutrophil immunity (adjusted P-value=1.95e-11 for all). In conclusion, DUOX2^+^ enterocytes have an active role in inflammation and are marked by the PSC risk gene HLA-DRB1 in PSC-IBD-I, suggesting that a part of its role in PSC-IBD pathogenesis is through antigen presentation (Supplemental results 2).

### Similar cell abundances between PSC-IBD and UC

Inflammation status predominately determines cellular composition of the biopsy. As depicted in Figure 1B and Figure 1C, the changes in cell population abundances were more pronounced in response to inflammatory conditions than variations between PSC-IBD and UC. In response to inflammation, the overall share of the epithelial compartment decreased, making room for an augmented immune compartment, while the stromal compartment remained relatively stable (Figure 1C). In general trends, both PSC-IBD and UC exhibited a decline in enterocyte subtypes during inflammation. Notably, DUOX2^+^ enterocytes showed a consistent increase in both diseases, reaching statistical significance in PSC-IBD ((Log-Ratio Value) LRV 0.8 at Credible Interval (CI) 0.95) and borderline significance in UC (LRV 0.9 at CI 0.90). Regarding immune compartment changes, both UC and PSC-IBD exhibited increases in CD8^+^IL17^+^ T cells during inflammation. B cells, Treg cells, and IgG plasma cells were significantly increased upon inflammation in both diseases. In PSC-IBD specifically, a rise in CD4^+^PD1^+^ T cells was observed (Supplemental results 3). Within the stromal compartment, inflammatory fibroblasts expanded during inflammation in both PSC-IBD (LRV 1.8 at CI 0.95) and UC (LRV 2.0 at CI 0.95). Conversely, WNT2B^+^ Fos-lo fibroblasts and microvascular cells exhibited decreases in both diseases (Supplemental results 3).

Interestingly, a significantly increased abundance of stem cells was noted in PSC-IBD compared to Healthy Controls (HC) (LRV 0.8 at CI 0.95) (Figure 1D and Supplemental Results 3).

### Differential expression analysis reveals differences between PSC-IBD and UC inflammation

Next, we studied the distinctive expression patterns that delineate inflammation in PSC-IBD and UC. Our approach involved conducting differential expression (DE) analyses for PSC-IBD-I versus PSC-IBD-NI and UC-I versus UC-NI across various cell types (Supplemental Results 4 and 5).

We found substantial differences in the number of DE genes per cell type between PSC-IBD and UC, as depicted in Figure 3B. Notably, the largest number of DE genes between in PSC-IBD-I and in PSC-IBD-NI was identified in inflammatory monocytes (n=407). In contrast, a distinct profile in inflammatory fibroblasts with a large number of DE genes was observed between UC-I and UC-NI (n=1058). Furthermore, two other types of fibroblasts (WNT2B^+^ FOS-lo 1 and WNT2B^+^ FOS-hi fibroblasts) showed large differential gene expression in UC-I vs UC-NI. In both PSC-IBD and UC, the DUOX2+ enterocyte and secretory TA cells emerged as consistently prominent cells with many DE genes between inflamed and non-inflamed. These results underscore the unique cellular signatures characterizing inflammation in PSC-IBD and UC, shedding light on distinct molecular mechanisms underlying these two inflammatory conditions.

**Figure 3:**
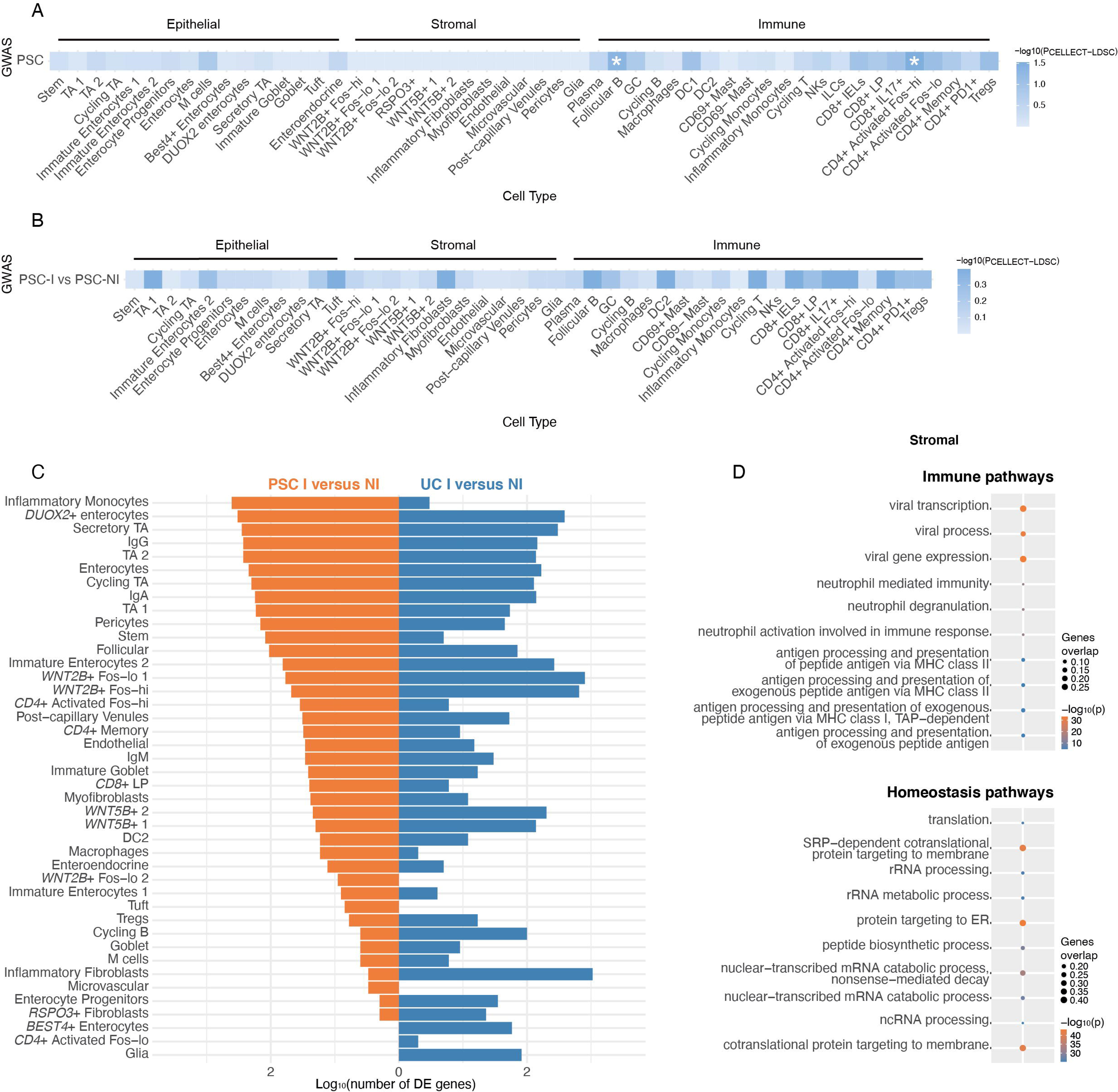
Follicular B and CD4+ Activated Fos-hi are enriched for heritable risk of PSC. Heatmap of partitioning heritability from GWAS of PSC into functional categories based on (A) PSC specifically expressed genes and (B) differentially expressed genes between PSC-I and PSC-NI states in each cell type. Categories reaching significance after Benjamini-Hochberg correction are denoted with an asterisk. (C) mirror barplot representing the number of differentially expressed genes on a log-scale per cell type for PSC-I compared to PSC-NI (orange) and UC-I compared to UC-NI (blue). (D) Differentially upregulated GO pathways in inflammatory monocytes in PSC-I versus PSC-NI. Pathways were manually divided into homeostasis and immune pathways. Color represents the log-normalized p-value of the pathway. Size represents the proportion of genes marking the pathways differentially upregulated in inflammatory monocytes in PSC-I as compared to PSC-NI. PSC, Primary Sclerosing Cholangitis; GWAS, genome-wide association study; PSC-I, PSC-inflammation; PSC-NI, PSC-non inflammation. GO, Gene Ontology

### Inflammatory monocytes are activated upon PSC-IBD, but not UC, inflammation

Intrigued by the vast transcriptomic changes displayed by inflammatory monocytes in PCS-IBD (as reflected by 407 DE genes), we further investigated their predicted functions within the context of PSC-IBD and UC inflammation (Supplemental results 6 and 7). Our dataset, comprising 137 PSC-IBD (69 inflamed and 68 non-inflamed) and 303 UC (257 inflamed and 46 non-inflamed) inflammatory monocytes, showed an increased abundance of this cell type upon inflammation in UC (LRV 1.23 CI 0.95), contrasting with the absence of a similar trend in PSC-IBD. Even while the absolute number of inflammatory monocytes is higher in UC samples, we find a marked difference in the number of DE genes between PSC-IBD-I versus PSC-IBD-NI (n=407) and UC-I versus UC-NI (n=3), which is unlikely to be attributable to a difference in statistical power.

The top pathways for PSC-IBD-I are housekeeping genes, however, ranking high also are many immune pathways with significant enrichment, including those associated with neutrophil immunity (adjusted p-value ∈ [5.09e-16, 7.81e-16]), proteolysis involved in cellular protein catabolic process (adjusted p-value = 1.94e-05), and antigen processing and presentation (adjusted p-value ∈ [2.65e-05, 2.69e-05]) (Figure 3C).

These findings illuminate the distinct molecular pathways associated with inflammatory monocytes in the context of PSC-IBD and UC inflammation, providing valuable insights into the nuanced immune responses characterizing these two conditions.

### No differential expression oncogenes and tumour suppressor genes in PSC-IBD-I compared to UC-I

We found an increased abundance of stem cells in PSC-IBD-NI, and not UC-NI, compared to HC, which may be linked to increased CRC risk in PSC-IBD. Therefore, we compared the expression of known cancer related genes (*i.e.* tumour suppressor and oncogenes) in stem cells between UC and PSC-IBD, in both inflamed and non-inflamed states[29]. For both the non-inflamed state and the inflamed state, we found that similar ratios of tumour suppressor to oncogenes were upregulated in stem cells in PSC-IBD and UC. Total numbers of stem cells in both PSC-IBD and UC were relatively low, which probably rendered the current analysis underpowered (Supplemental results 9).

### Cell-cell interactions show antigen presentation through inflammatory monocytes in PSC-IBD

To unravel the intricate landscape of cell-cell interactions in the context of PSC-IBD inflammation, we focused on the cell types that showed most differentially expressed genes upon inflammation —namely, DUOX2+ enterocytes and inflammatory monocytes (Figure 3C). These cell types were deemed central to the inflammatory process. Furthermore, since two previously published studies[30,31] and our data suggest a central role for antigen presentation in the pathogenesis of PSC, we focused the analysis on interactions through MHC-II encoding HLA 2 genes (Supplemental results 10).

Our analysis extended to exploring monocyte interactions with the rest of the immune cells. Remarkably, the majority of cell-cell interactions during inflammation were shared between PSC-IBD and UC. In both conditions, inflammatory monocytes and DUOX2^+^ enterocytes were predicted to communicate with innate (macrophages and dendritic cells), as well as adaptive immune cells (Tregs, cycling T cells, and CD4^+^PD1^+^ T cells) (Figure 4B).

**Figure 4:**
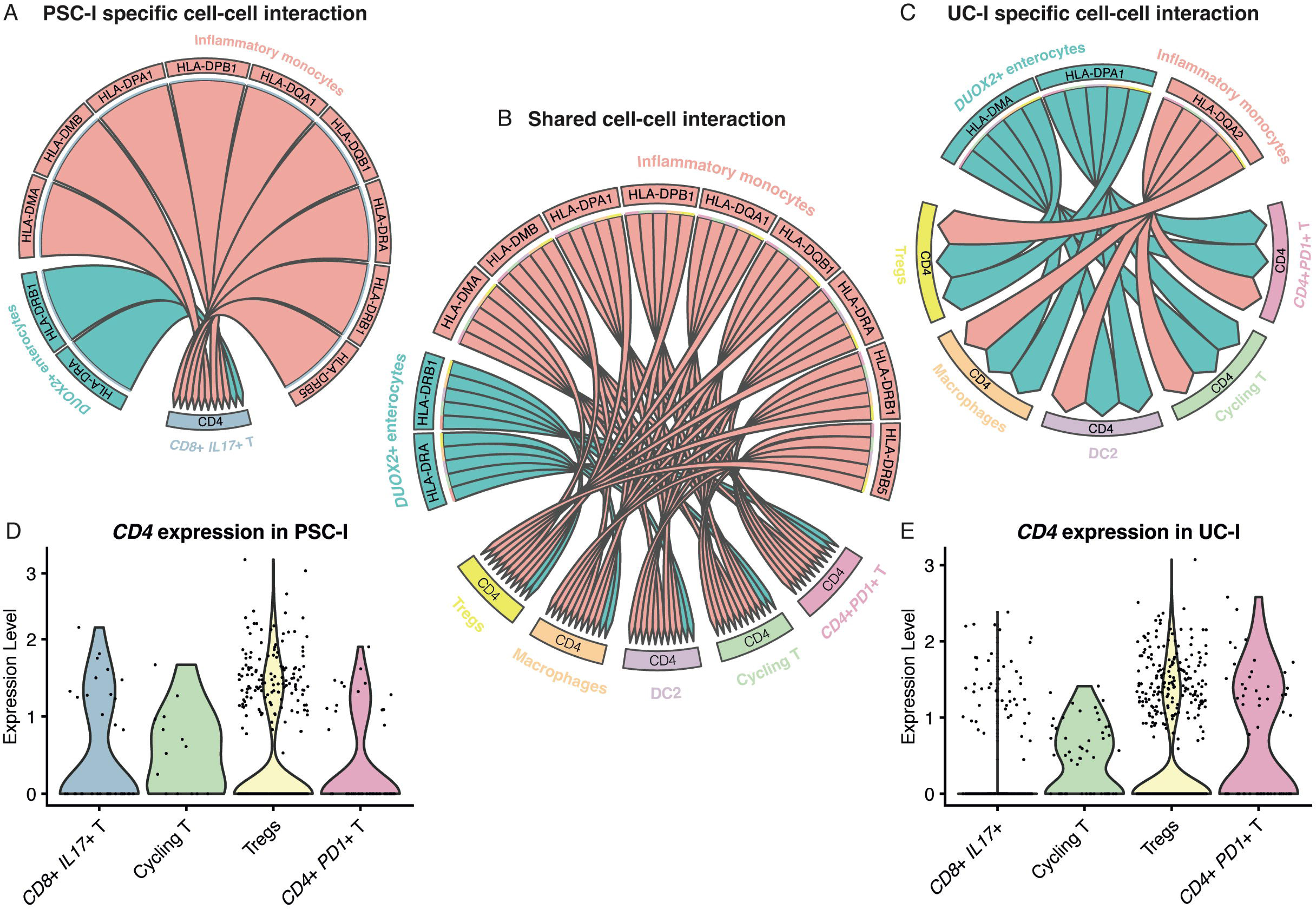
A: circos plot representing the unique MHC-II cell-cell interactions for inflammatory monocytes and DUOX2+ enterocytes as senders observed in PSC-I samples. B: circos plot representing the shared MHC-II cell-cell interactions for inflammatory monocytes and DUOX2+ enterocytes as senders observed in PSC-I and UC-I samples. C: circos plot representing the unique MHC-II cell-cell interactions for inflammatory monocytes and DUOX2+ enterocytes as senders observed in UC-I samples. D: Expression level of CD4 in different T cell subtypes in PSC-I samples E: Expression level of CD4 in different T cell subtypes in UC-I samples

However, a distinctive feature in PSC-IBD was the communication between both inflammatory monocytes and DUOX2^+^ enterocytes with CD8^+^IL17^+^ T cells through the CD4 coreceptor (Figure 4A). This finding is particularly noteworthy since CD8^+^IL17^+^ T cells are conventionally recognized as the central inflammatory cells in UC, with this cell-cell interaction being absent in our UC analysis (Figure 4C). Furthermore, in PSC-IBD, the communication of these cells through CD4 suggests that CD8^+^IL17^+^ T cells are CD4^+^CD8^+^ double positive T cells. Indeed, CD8^+^IL17^+^ T cells in PSC-IBD appear to express more CD4 compared to CD8^+^IL17^+^ T cells in UC (Figure 4D and Figure 4E), aligning with the findings of Shaw et al, who identified CD4+IL17+ T cells as the central inflammatory cell in PSC-IBD[31]. This observation provides a nuanced understanding of the immune cell interactions specific to PSC-IBD inflammation.

### Heritability analysis and PSC risk genes exhibit distinct expression patterns in PSC-IBD inflammation

To explore the impact of established PSC risk genes on PSC-IBD, we leveraged data from the latest PSC genome-wide association study and its meta-analysis[26,32]. Our objective was to identify the cell types in PSC-IBD that carry the highest burden of known PSC risk genes and are therefore potentially crucial in PSC-IBD pathogenesis. Using a heritability analysis, correcting for the number of genes expressed by each cell and number of cells per cell types, we found that follicular B cells and CD4+PD1+ T cells express the highest number of PSC risk genes, as depicted in Figure 3A. When trying to identify which cell type carries the highest known PSC risk gene burden in PSC-IBD I vs NI, we found no significant results, potentially due to a lack of power (Figure 3A and Supplemental results 8).

When looking at differential gene expression of the known PSC risk genes in PSC-IBD-I vs PSC-IBD-NI, we found that the known PSC risk genes might play a role in cells other than adaptive immune cells during PSC-IBD inflammation (Supplementary Figure 2). In the epithelial compartment, particularly in tuft cells and enteroendocrine cells, expression of PSC risk genes was observed in PSC-IBD during inflammation. Notably, PSC risk genes such as *FUT2*, *GPR35*, *PRDX5*, and *PSMG1* exhibited expression across multiple cell types within the epithelial compartment during inflammation. In the mesenchymal compartment, post-capillary venules, endothelial cells, and microvascular cells demonstrated expression of PSC risk genes in PSC-IBD during inflammation. Specifically, PSC risk genes including *ATXN2*, *HDAC7*, *NKX2-3*, *RIC8B*, *STRN4*, and *TCF4* were found to be expressed across various cell types within the mesenchymal compartment upon inflammation (Supplementary Figure 2).

Intriguingly, when assessing the expression of known PSC risk genes, we observed that several of these genes—namely *ATXN2*, *FUT2*, *GPR35*, *NKX2-3*, and *TCF4*—exhibited minimal differential expression in the immune compartment during inflammation. This finding suggests a distinctive pattern in the regulation of these genes in immune cells compared to their more pronounced expression in epithelial and stromal cells during PSC-IBD inflammation (Supplementary Figure 2).

## Discussion

Here, we present our study on whole-biopsy single-cell gene expression in PSC-IBD. Our data strongly support the hypothesis that a distinct pathomechanism fuels inflammation in PSC-IBD compared to UC. Crucially, our study underscores the pivotal role of inflammatory monocytes as key orchestrators of inflammation in PSC-IBD. These cells emerge as central players in driving the inflammatory cascade, shedding light on a previously underappreciated aspect of PSC-IBD pathogenesis. By elucidating these intricate cellular mechanisms, our study not only enhances our understanding of PSC-IBD pathophysiology but also offers valuable insights into potential therapeutic targets. In addition, the identification of inflammatory monocytes and DUOX2^+^ enterocytes as key drivers of inflammation open avenues for targeted interventions aimed at modulating these specific cell populations, thereby offering prospects for more effective and tailored therapeutic strategies in PSC-IBD management.

Our study reveals a notable shift in cell type proportions upon inflammation in PSC-IBD, mirroring trends observed in UC. Specifically, we observed a decrease in BEST4^+^ enterocytes and an expansion of inflammatory fibroblasts, consistent with previous findings in UC[20]. Furthermore, we noted an increased abundance of IgG plasma cells during inflammation, a phenomenon documented in both PSC-IBD and UC[31,33].

Our research reveals a novel subtype of enterocytes, DUOX2^+^ enterocytes, identified by the expression of *DUOX2* and *LCN2,* which are prominently enriched in inflamed colonic regions compared to non-inflamed colonic regions, particularly in PSC-IBD. DUOX2 and LCN2 have been associated with innate immune responses and microbiota regulation in IBD[34–38]. A recent publication has described this cell type as well in the context of UC[39]. This discovery suggests a fundamental involvement of DUOX2^+^ enterocytes in the inflammatory process, potentially mediated through major histocompatibility complex class II (MHC II) molecule expression. While epithelial cells expressing MHC II during inflammation have been documented previously, our study is among the first to identify this specific cell subtype with that function[40]. Interestingly, recent research implicates DUOX2-expressing epithelial cells in Crohn’s disease inflammation, further supporting the significance of our findings[41]. Our study unveils marker pathways for DUOX2^+^ cells primarily involved in immune responses and antigen presentation, a novel finding in the context of PSC-IBD.

Distinguishing expression profiles and functional roles driving inflammation in PSC-IBD compared to UC are evident. While UC primarily involves substantial changes in inflammatory fibroblasts, PSC-IBD inflammation predominantly activates inflammatory monocytes. Genomic studies associating PSC-IBD with variants at the *HLA-DRB1* locus suggest an autoimmune pattern akin to celiac disease, where unknown antigens trigger inflammation[12]. Inflammatory monocytes may play a crucial role in presenting these antigens and instigate inflammation, although further functional studies are warranted for validation.

The increased risk of colorectal cancer (CRC) in PSC-IBD patients underscores a distinct pathophysiology compared to UC. Our study, along with recent research, suggests a distinct role for CD4+IL17+FOXP3+ T cells in PSC-IBD dysplasia progression[31]. We observed increased communication to CD8^+^IL17^+^ T cells in PSC-IBD inflammation, most likely involving CD4^+^CD8^+^ T cells. Additionally, our data suggest an increased abundance of stem cells in PSC-IBD, possibly contributing to CRC risk[42]. Our study was underpowered to confirm that our CD8+CD4+IL17+ T cells are the same cells as the previously published CD4+IL17+FOXP3+ T cells.

Despite limitations such as sample size constraints and separate sampling of inflamed and non-inflamed tissues, our study lays a solid foundation for comprehending the intricate cellular landscape in PSC-IBD. Functional validation and further research are imperative to elucidate the mechanisms underlying the observed gene expression differences and their clinical implications.

## Supporting information

Full Supplementary Data

## Author contributions to the manuscript

A.B., P.P, S.S., J.B. and W.T.C.U.V. participated in analyses; E.B., S.d.J., M.G.P.v.d.W. and W.T.C.U.V. participated in lab work. All authors participated in discussion of the results and writing and editing of the manuscript.

## Data availability

Data will be made available upon request through the EGA server under nr XXX. Scripts may be found here https://github.com/WeersmaLabIBD/PSC_project.

## Acknowledgments

We would like to thank all the IBD patients who participate in the Dutch IBD Biobank. This nationwide PSI project is part of and funded by the Netherlands Federation of University Medical Centers and has received initial funding from the Dutch Government (from 2007-2011). The PSI currently facilitates the uniform nationwide collection of information on and biomaterials of thirteen other diseases.

## Abbreviations used in this manuscript

IBD: Inflammatory Bowel Disease
PSC: Primary Sclerosing Cholangitis
PDC-IBD: Primary sclerosing cholangitis associated Inflammatory bowel disease
ScRNAseq: single-cell messenger RNA sequencing
UC: Ulcerative colitis
I and NI: Inflamed and Non-Inflamed
CRC: Colorectal carcinoma
CD: Crohn’s disease
anti-TNF: anti-Tumour Necrosis Factor Alpha
HC: Healthy control
IRB: Institutional Review Board
FCS: Foetal Calf Serum
PBS: Phosphate buffered saline
QC: Quality control
PCA: Principal Component Analysis
DAB: 3,3′-Diaminobenzidine
NDP: NanoZoomer Digital Pathology

## Bibliography

1 Bambha K, Kim WR, Talwalkar J, et al. Incidence, clinical spectrum, and outcomes of primary sclerosing cholangitis in a United States community. Gastroenterology. 2003;125:1364–9. doi: 10.1016/j.gastro.2003.07.011

2 Liang H, Manne S, Shick J, et al. Incidence, prevalence, and natural history of primary sclerosing cholangitis in the United Kingdom. Medicine (Baltimore). 2017;96:e7116. doi: 10.1097/MD.0000000000007116

3 Toy E, Balasubramanian S, Selmi C, et al. The prevalence, incidence and natural history of primary sclerosing cholangitis in an ethnically diverse population. BMC Gastroenterol. 2011;11:83. doi: 10.1186/1471-230X-11-83

4 Boonstra K, Weersma RK, van Erpecum KJ, et al. Population-based epidemiology, malignancy risk, and outcome of primary sclerosing cholangitis. Hepatology. 2013;58:2045–55. doi: 10.1002/hep.26565

5 Trivedi PJ, Bowlus CL, Yimam KK, et al. Epidemiology, Natural History, and Outcomes of Primary Sclerosing Cholangitis: A Systematic Review of Population-based Studies. Clin Gastroenterol Hepatol. 2022;20:1687–1700.e4. doi: 10.1016/j.cgh.2021.08.039

6 van Munster KN, Mol B, Goet JC, et al. Disease burden in primary sclerosing cholangitis in the Netherlands: A long-term follow-up study. Liver Int. 2023;43:639–48. doi: 10.1111/liv.15471

7 Lakatos P-L, Lakatos L. Risk for colorectal cancer in ulcerative colitis: changes, causes and management strategies. World J Gastroenterol. 2008;14:3937–47. doi: 10.3748/wjg.14.3937

8 Chapman MH, Thorburn D, Hirschfield GM, et al. British Society of Gastroenterology and UK-PSC guidelines for the diagnosis and management of primary sclerosing cholangitis. Gut. 2019;68:1356–78. doi: 10.1136/gutjnl-2018-317993

9 Olén O, Erichsen R, Sachs MC, et al. Colorectal cancer in Crohn’s disease: a Scandinavian population-based cohort study. Lancet Gastroenterol Hepatol. 2020;5:475–84. doi: 10.1016/S2468-1253(20)30005-4

10 de Vries AB, Janse M, Blokzijl H, et al. Distinctive inflammatory bowel disease phenotype in primary sclerosing cholangitis. World J Gastroenterol. 2015;21:1956–71. doi: 10.3748/wjg.v21.i6.1956

11 Hedin CRH, Sado G, Ndegwa N, et al. Effects of Tumor Necrosis Factor Antagonists in Patients With Primary Sclerosing Cholangitis. Clin Gastroenterol Hepatol. 2020;18:2295–2304.e2. doi: 10.1016/j.cgh.2020.02.014

12 Jiang X, Karlsen TH. Genetics of primary sclerosing cholangitis and pathophysiological implications. Nat Rev Gastroenterol Hepatol. 2017;14:279–95. doi: 10.1038/nrgastro.2016.154

13 Nordenvall C, Olén O, Nilsson PJ, et al. Colectomy prior to diagnosis of primary sclerosing cholangitis is associated with improved prognosis in a nationwide cohort study of 2594 PSC-IBD patients. Aliment Pharmacol Ther. 2018;47:238–45. doi: 10.1111/apt.14393

14 Konnikova L, Boschetti G, Rahman A, et al. High-dimensional immune phenotyping and transcriptional analyses reveal robust recovery of viable human immune and epithelial cells from frozen gastrointestinal tissue. Mucosal Immunol. 2018;11:1684–93. doi: 10.1038/s41385-018-0047-y

15 Uniken Venema WTC, Ramírez-Sánchez AD, Bigaeva E, et al. Gut mucosa dissociation protocols influence cell type proportions and single-cell gene expression levels. Sci Rep. 2022;12:9897. doi: 10.1038/s41598-022-13812-y

16 Anon. Protocol - TotalSeqTM-A Antibodies and Cell Hashing with 10x Single Cell 3’ Reagent Kit v3 3.1 Protocol. Available at: https://www.biolegend.com/en-us/protocols/totalseq-a-antibodies-and-cell-hashing-with-10x-single-cell-3-reagent-kit-v3-3-1-protocol.

17 Anon. Single Cell Gene Expression - 10x Genomics. Available at: https://www.10xgenomics.com/products/single-cell-gene-expression.

18 Heaton H, Talman AM, Knights A, et al. Souporcell: robust clustering of single-cell RNA-seq data by genotype without reference genotypes. Nat Methods. 2020;17:615–20. doi: 10.1038/s41592-020-0820-1

19 Satija R, Farrell JA, Gennert D, et al. Spatial reconstruction of single-cell gene expression data. Nat Biotechnol. 2015;33:495–502. doi: 10.1038/nbt.3192

20 Smillie CS, Biton M, Ordovas-Montanes J, et al. Intra- and Inter-cellular Rewiring of the Human Colon during Ulcerative Colitis. Cell. 2019;178:714–730.e22. doi: 10.1016/j.cell.2019.06.029

21 Silverman JD, Roche K, Holmes ZC, et al. Bayesian Multinomial Logistic Normal Models through Marginally Latent Matrix-T Processes. Journal of Machine Learning Research. 2022;23:1–42.

22 Finucane HK, Reshef YA, Anttila V, et al. Heritability enrichment of specifically expressed genes identifies disease-relevant tissues and cell types. Nat Genet. 2018;50:621–9. doi: 10.1038/s41588-018-0081-4

23 Bailey MH, Tokheim C, Porta-Pardo E, et al. Comprehensive Characterization of Cancer Driver Genes and Mutations. Cell. 2018;173:371–385.e18. doi: 10.1016/j.cell.2018.02.060

24 Jin S, Guerrero-Juarez CF, Zhang L, et al. Inference and analysis of cell-cell communication using CellChat. Nat Commun. 2021;12:1088. doi: 10.1038/s41467-021-21246-9

25 Timshel PN, Thompson JJ, Pers TH. Genetic mapping of etiologic brain cell types for obesity. Elife. 2020;9:e55851. doi: 10.7554/eLife.55851

26 Ji S-G, Juran BD, Mucha S, et al. Genome-wide association study of primary sclerosing cholangitis identifies new risk loci and quantifies the genetic relationship with inflammatory bowel disease. Nat Genet. 2017;49:269–73. doi: 10.1038/ng.3745

27 Finak G, McDavid A, Yajima M, et al. MAST: a flexible statistical framework for assessing transcriptional changes and characterizing heterogeneity in single-cell RNA sequencing data. Genome Biol. 2015;16:278. doi: 10.1186/s13059-015-0844-5

28 Zimmerman KD, Espeland MA, Langefeld CD. A practical solution to pseudoreplication bias in single-cell studies. Nat Commun. 2021;12:738. doi: 10.1038/s41467-021-21038-1

29 Palaniappan A, Muthamilselvan S, Sarathi A. COADREADx: A comprehensive algorithmic dissection unravels salient biomarkers and actionable insights into the discrete progression of colorectal cancer. 2024;2022.08.16.22278877.

30 Cargill T, Culver EL. The Role of B Cells and B Cell Therapies in Immune-Mediated Liver Diseases. Front Immunol. 2021;12:661196. doi: 10.3389/fimmu.2021.661196

31 Shaw DG, Aguirre-Gamboa R, Vieira MC, et al. Antigen-driven colonic inflammation is associated with development of dysplasia in primary sclerosing cholangitis. Nat Med. 2023;29:1520–9. doi: 10.1038/s41591-023-02372-x

32 Alberts R, de Vries EMG, Goode EC, et al. Genetic association analysis identifies variants associated with disease progression in primary sclerosing cholangitis. Gut. 2018;67:1517–24. doi: 10.1136/gutjnl-2016-313598

33 Uzzan M, Martin JC, Mesin L, et al. Ulcerative colitis is characterized by a plasmablast-skewed humoral response associated with disease activity. Nat Med. 2022;28:766–79. doi: 10.1038/s41591-022-01680-y

34 Makhezer N, Ben Khemis M, Liu D, et al. NOX1-derived ROS drive the expression of Lipocalin-2 in colonic epithelial cells in inflammatory conditions. Mucosal Immunol. 2019;12:117–31. doi: 10.1038/s41385-018-0086-4

35 Haberman Y, Tickle TL, Dexheimer PJ, et al. Pediatric Crohn disease patients exhibit specific ileal transcriptome and microbiome signature. J Clin Invest. 2014;124:3617–33. doi: 10.1172/JCI75436

36 Grasberger H, Magis AT, Sheng E, et al. DUOX2 variants associate with preclinical disturbances in microbiota-immune homeostasis and increased inflammatory bowel disease risk. J Clin Invest. 2021;131:e141676, 141676. doi: 10.1172/JCI141676

37 Burgueño JF, Fritsch J, González EE, et al. Epithelial TLR4 Signaling Activates DUOX2 to Induce Microbiota-Driven Tumorigenesis. Gastroenterology. 2021;160:797–808.e6. doi: 10.1053/j.gastro.2020.10.031

38 Singh V, Yeoh BS, Chassaing B, et al. Microbiota-inducible Innate Immune, Siderophore Binding Protein Lipocalin 2 is Critical for Intestinal Homeostasis. Cell Mol Gastroenterol Hepatol. 2016;2:482–498.e6. doi: 10.1016/j.jcmgh.2016.03.007

39 MacFie TS, Poulsom R, Parker A, et al. DUOX2 and DUOXA2 form the predominant enzyme system capable of producing the reactive oxygen species H2O2 in active ulcerative colitis and are modulated by 5-aminosalicylic acid. Inflamm Bowel Dis. 2014;20:514–24. doi: 10.1097/01.MIB.0000442012.45038.0e

40 Heuberger CE, Janney A, Ilott N, et al. MHC class II antigen presentation by intestinal epithelial cells fine-tunes bacteria-reactive CD4 T-cell responses. Mucosal Immunol. 2023;S1933–0219(23)00032–6. doi: 10.1016/j.mucimm.2023.05.001

41 Li J, Simmons AJ, Chiron S, et al. A Specialized Epithelial Cell Type Regulating Mucosal Immunity and Driving Human Crohn’s Disease. bioRxiv. 2023;2023.09.30.560293. doi: 10.1101/2023.09.30.560293

42 Walcher L, Kistenmacher A-K, Suo H, et al. Cancer Stem Cells-Origins and Biomarkers: Perspectives for Targeted Personalized Therapies. Front Immunol. 2020;11:1280. doi: 10.3389/fimmu.2020.01280

